# Medical, socioeconomic, and geographic disparities in primary health care access and utilization: A population-based study of 8038 individuals aged one year and older in rural Uganda

**DOI:** 10.1101/2025.10.23.25338656

**Authors:** Saadiyah Mayet, Christin Puthur, Betty Nabatte, Benjamin Tinkitina, Christopher K Opio, Yin-Cong Zhi, Narcis B Kabatereine, Goylette F Chami

**Author notes:** These authors contributed equally to this work (Saadiyah Mayet, Christin Puthur).

## Abstract

**Background:** There is limited data on trends in primary health care (PHC) access and utilisation in rural sub-Saharan Africa, with a particular lack of knowledge on disparities beyond geographic barriers.

**Methods:** A cross-sectional study was conducted within the community-based cohort, SchistoTrack. 2191 households were randomly sampled from 52 rural villages in Pakwach, Buliisa, and Mayuge Districts in Uganda. All household members aged one year and older were surveyed between 2022-2024, resulting in 8038 individuals with baseline health access information. Key outcomes included any care sought within the month preceding the study, type of care sought, and usual source of care. Using logistic regressions with standard errors clustered by household and village, we examined a comprehensive and diverse set of medical, socioeconomic, and geographic determinants.

**Results:** Only 8.2% (659/8038) of participants sought any care, with only 67.5% (445/659) of those participants seeking PHC. Having current symptoms, a previously diagnosed disease, older age, and being female were positively associated (Odds ratios (ORs) 1.02-2.85) with higher odds of seeking any care in the past month. Among participants who sought care, living in a Western district (Pakwach or Buliisa) was associated with higher odds (ORs 9.40-10.77) of seeking care from PHC centres than living in the Eastern study district (Mayuge). Current alcohol use among adults aged 20 years and older was associated with lower odds of seeking PHC (OR 0.43). Distance to a PHC centre and living in the Western districts were negatively (OR 0.72) and positively associated (ORs 4.70-5.64) with having PHC as the usual source of care for the household.

**Conclusions:** Medical and demographic factors influenced the decision to seek any care, but PHC usage was largely determined by regional and spatial disparities suggesting different avenues for targeted interventions seeking to increase general health access vs PHC usage.

## Introduction

The lowest indicators of universal health coverage (UHC) are mostly found in sub-Saharan Africa (SSA) [1]. Improving the access, quality, and financing of primary health care (PHC) services determines progress towards UHC principles of inclusivity and affordability of essential services for a wide range of common diseases [2]. Yet, there is limited data on PHC access and utilisation for individuals in rural SSA that incorporates the diverse range of biomedical, socioeconomic, and spatial influences.

Less than 50% of individuals with a usual source of care access public sector care at the primary level [2]. PHC depends on population-based platforms for care delivery, including lower tier, unspecialised health centres that are the first point of contact for patients [3]. Access to PHC—one component of the choice of usual care—has primarily relied on studies that focus on geographic barriers, primarily guided by large-scale mapping of distances to health centres [4] or identification of rough terrain and poor road infrastructure [5] [6] [4] [7]. With respect to the utilisation of PHC in SSA, women surprisingly, despite being responsible for the care of young children and having access to PHC through antenatal and maternal health services, have been suggested to be less likely to seek care [8]. Age has been reported as an important factor in mostly descriptive studies [9] [10], which does not allow separation from other confounders such as geographic barriers to access. Higher educational status and literacy often have been associated with a higher likelihood of seeking care [10] [7]. Though less studied, symptom type and perceived severity of illness may also affect the decision to seek care as well as the type of care [11] [10]. Cost of care to the patient also has been commonly reported as a barrier to care-seeking [5] [11] [10] with suggestions that higher income is associated with seeking public or private care as opposed to other avenues such as traditional medicine [12]. In cities of SSA, it has been shown that individuals have plentiful access to PHC and the agency to choose providers [13], but it is unclear whether this finding generalises to rural areas with poorer health infrastructure. Overall, it remains an open question as to the relative contribution of the diverse factors that influence PHC usage to understand whom to target for improving UHC, given a conventional focus on limited determinants of PHC usage or dimensions of UHC. A broader scope and more comprehensive set of dimensions within a single study is needed to understand what disparities exist within rural communities. Moreover, patient priorities and medical needs are poorly understood. There is a need for studies that incorporate the health knowledge a patient has of their own medical history, as well as the knowledge a patient has of the medical histories of other household (HH) members.

We conducted a population-based study to predict PHC usage for individuals aged one year and older from 52 rural Ugandan communities that were part of the SchistoTrack cohort. By assessing the contribution of patient health knowledge of medical histories and symptoms as well as HH distance to PHC centres, water, sanitation, and hygiene (WASH), costs of care, and socio-demographic indicators, we sought to address the following questions. What are the determinants of seeking care and the usual source of care? Do disparities exist and which dimensions beyond geographic barriers most influence PHC usage?

## Methods

### Study context

This study was conducted in Mayuge, Buliisa, and Pakwach Districts in Uganda along Lake Victoria, Lake Albert, and the Nile River, respectively. According to the 2024 census, Buliisa, Mayuge, and Pakwach had populations of 167894, 577563, and 206961, respectively [14]. Public, government-funded care has been reported to account for 66% of health care delivery in Uganda [15]. Trained health workers, whether nurses, medical officers, or doctors, are supported by laymen as part of village health teams (VHTs) (the lowest cadre of health care workers), which are especially important in rural areas of Uganda. There are an estimated 5.17 VHT members per 1000 individuals compared to 0.03 physicians and 0.46 nurses per 1000 individuals [16] [17] [18]. Private drug shops that are informally established within villages or trading centres across villages also play a large role in health care delivery in rural Uganda and elsewhere in SSA as the main avenue of informal care [19].

### Participant sampling

Participants were recruited as part of the SchistoTrack study [20] [21], which is a prospective, community-based cohort set up to examine the development and progression of liver fibrosis related to schistosomiasis. Three rounds of recruitment from 2022 to 2024 were completed during January to February of each year. Approximately 40 HHs were systematically randomly sampled using village registers ordered by year of first residence from 52 villages; details are provided elsewhere [22]. Within the 2191 randomly sampled HHs, information was collected through direct interviews with the HH head and wife or husband of the HH head on every person aged one year and older in the home. A total of 8042 individuals were surveyed. An electronic pre-validated survey programmed in Open Data Kit (ODK) Collect v2022.4.2 in 2022, and ODK Collect v2023.2.4 in 2023 and 2024 was used [20].

### Outcomes

Health access and utilisation outcomes of any care sought, and the main type of care sought within the one month preceding study recruitment, were coded as binary indicators and measured at the individual level. Any care sought was equal to one if a participant had sought medical care regardless of the reason. The type of care sought was coded as one if a participant sought public care either from formal PHC centres (government only) or from VHTs (lay health workers within each village), and otherwise zero to represent private care. PHC included government health centres that were not specialist facilities and not hospitals (at any level where the lowest was regional). VHTs are comprised of government lay health workers at the lowest tier of the health system, more akin to informal or community-based public health care. Private care included private health clinics, local medicine shops within villages and at trading centres serving multiple villages, traditional healers, herbalists, traditional birth attendants, bone setters, spiritual counsellors, witch doctors, or any other category of care or where the type of care sought was unknown. At the HH level, the usual care sought—without restriction to a recall period— was assessed and coded as binary where the outcome was equal to one if the HH respondent indicated that the main type of care used was PHC centres (government only) or VHTs, and otherwise, zero. This definition aligns with the World Health Organization and UNICEF operational framework for primary health care, which emphasises that PHC should be the first point of contact for people seeking care, government-led, and integrated with other health services [23]. It is also consistent with the global consensus that PHC must include population and community-based care, as outlined in the Alma-Ata Declaration [3]. Henceforth, PHC will be used to refer to government-provided care, inclusive of VHTs unless otherwise specified.

### Biomedical, socioeconomic, and spatial covariates

To assess patient-prioritised medical needs, medical histories with no restricted timeline and clinical symptoms experienced in the month preceding recruitment were collected within the HH survey. Information was collected on self-reported past diagnoses for both infectious diseases (IDs) using validated modules from World Health Surveys [24] [25], and for non-communicable diseases (NCDs) using modules on hypertension, stroke, cholesterol, and diabetes from World Health Organization (WHO) STEPwise approach (STEPS) surveys [26]. Using a pre-defined list of IDs and NCDs informed by previous studies [24] (Supplementary Table S1), self-reported medical histories were elicited by asking respondents about every individual within the HH and whether they were told by a doctor or health worker as having each disease at any time in the past. Current alcohol consumption and smoking within the past 12 months preceding the study were collected for participants aged ≥10 years based on modules from the STEPS surveys and coded as binary variables. NCDs, including raised blood pressure, raised cholesterol, raised blood sugar, and heart disease, were only asked about within the HH survey for participants aged ≥20 years, and sexually transmitted infections (STIs) were only asked for participants aged ≥10 years. In addition to specific disease indicators, binary indicators were calculated based on whether the person of interest reported a history of any IDs as well as whether anyone else in the HH of the participant of interest had reported at least one ID, including all possible IDs except STIs and HIV. For adult-only sub-group analyses, variables on current alcohol consumption and smoking were added to the candidate variables, and all ID indicators were broadened to include STIs and NCD indicators broadened to include NCDs collected only for participants aged ≥20.

Clinical symptoms were recorded as free text and grouped following the procedure described in Chami and colleagues [24]. A binary indicator was generated that was equal to one if a participant had experienced any symptoms in the month preceding recruitment into the cohort. Binary indicators for specific symptoms of fever and diarrhoea that served as flagship symptoms [24] for several diseases were also constructed. Participants were asked whether they had taken any Western medicines in the past month, and if so asked which medicine was taken among options of praziquantel, other deworming medicines, antimalarials, antibiotics, anti-retroviral therapies (ARTs), blood pressure medication, paracetamol, aspirin, ibuprofen, and blood thinners, with an option for other. ARTs, blood pressure medicine, and blood thinners were taken as indicators of chronic conditions and a separate binary indicator equal to one was constructed if the participant had taken any of these medicines within the past month with all other responses (no medicines or other medicines) coded as zero.

Beyond individual-level biomedical indicators, indicators related to other HH members were recorded. These indicators included binary variables of whether any other HH member had an ID or NCD, as previously described. A binary indicator of whether there were any deaths in the HH within the past three years was also recorded. These indicators were included to capture the influence of health knowledge from learning about diseases from other individuals and any direct familial influences on health care seeking.

Demographic, socioeconomic, and water, sanitation, and hygiene (WASH) data were collected at the individual or HH level. Participant age, gender, occupation, tribe, religion, educational attainment, HH social status, number of individuals in the HH, years of HH settlement in the village, home ownership, number of rooms in the house, home quality, and HH WASH were considered. For tribe and religion, these variables were examined and coded as majority tribe and majority religion to represent whether a participant belonged to the majority tribe or majority religion of their village to understand the extent a participant belonged to a minority group. Detailed variable definitions are provided elsewhere [20].

For spatial factors, the minimum distances from HHs to drug shops and government health centres were calculated by computing the Euclidean distance between coordinate points on the World Geodetic System (WGS) 84 ellipsoid. Waypoints of homes and health infrastructure were recorded at the time of the HH survey within ODK Collect with an accuracy range of 5-10 metres on tablets using Android Pie 9. Only local medicine shops listed by village leaders and VHT members as currently used by the participants were visited, and only those that supplied medicines were included in analyses. For local medicine shops and PHC centres, respectively, in each district, there were 13 and three in Pakwach, nine and three in Buliisa, and 20 and four in Mayuge functioning facilities at the time of survey. A categorical variable of the study district was also included to capture elements of the study design (sampling within districts) and broader district-level differences in health care infrastructure [27]. To account for temporal differences at the point of recruitment, the year of recruitment was included as a categorical variable.

### PHC centre variables

Travel time was recorded for participants who had visited a government PHC centre, using categories of within 10 minutes, within 30 minutes, within 60 minutes, more than one hour, more than two hours, and unknown. All participants who sought care were asked to report the cost of care received, excluding travel costs, in Ugandan shillings (UGX). Participants who had visited a government health centre in the past month were also asked to share the final diagnoses given by the health care worker, and these responses were entered as free text.

### Statistical analysis

All analyses were conducted in R v4.2.1. 8038 participants surveyed had complete data for all outcomes and covariates and were the focus of all analyses, with two adults missing STIs and NCDs, and two participants with incomplete HH information. Multilevel logistic regression was run for the individual-level outcomes of any care sought with random effects for HH and village. Intraclass clustering coefficients were calculated for empty and adjusted multilevel models. Ordinary logistic regression with clustered standard errors at the HH level [28] was run for the outcome of the type of care which was a sub-group analysis of only participants who had sought care. To assess the addition of covariates for STIs and NCDs for adults ≥20 years, both individual-level models were rerun using ordinary logistic regression, with clustered standard errors at the HH level. For the usual source of care, logistic regressions were run at the HH level using only covariates measured at the HH level apart from recalculated medical history indicators as follows. To focus on how medical histories within the HH influence the usual source of care for the HH, IDs (including STIs) and NCD indicators were recalculated and equal to one if anyone in the home had a disease in that category. Covariate selection was done for each model by finding the best fit parsimonious model, using backward selection and the lowest Bayesian Information Criterion (BIC) [29]. For regression models where the year of recruitment variable was significant, floating absolute risks (FARs) were computed with adapted code from the Epi package in R [30] to test the choice of reference category. Each model was checked for multicollinearity by calculating the variance inflation factors (VIFs) [31]. Ten-fold cross-validated (CV) [32] areas under the receiver operating characteristic curves (AUCs) were calculated for all models.

## Results

### Perceived illness and PHC use across districts

Among 8038 participants recruited in SchistoTrack, 43.9% (3532/8038) reported one or more past IDs, and 15.2% (1220/8038) reported one or more past NCDs. The most common past ID and NCD were uncomplicated malaria and eczema, respectively (Table 1, Supplementary table S1). There were 3.5% (285/8038) participants who reported one or more symptoms in the month before recruitment, of whom 5.6% (16/285) reported fever and 9.8% (28/285) diarrhoea. The percentage of participants who reported a symptom in the past month was 6.8% (224/3313) in Pakwach, 2% (50/2455) in Buliisa, and 0.5% (11/2270) in Mayuge (χ^2^ = 178.7, p< 0.001). Among those who reported past symptoms, 4.9% (11/224) in Packwach and 10% (5/50) in Buliisa reported fever, with none reported in Mayuge (Fisher’s exact test, p = 0.273), and 8.9% (20/224) in Packwach and 14% (7/50) in Buliisa reported diarrhoea, with none reported in Mayuge (Fisher’s exact test, p = 0.365).

**Table 1.** Participant characteristics.

|  |  | Total<br>(N=8038) | Care seeking<br>(N=659) | Not care seeking<br>(N=7379) |
| --- | --- | --- | --- | --- |
| **BIOMEDICAL FACTORS** |  |  |  |  |
| Any symptoms |  | 285 (3.5%) | 66 (10.0%) | 219 (3.0%) |
| IDs for other HH members |  | 4942 (61.5%) | 479 (72.7%) | 4463 (60.5%) |
| NCDs for other HH members |  | 2230 (27.7%) | 208 (31.6%) | 2022 (27.4%) |
| History of IDs |  | 3532 (43.9%) | 423 (64.2%) | 3109 (42.1%) |
| History of NCDs |  | 1220 (15.2%) | 131 (19.9%) | 1089 (14.8%) |
| History of any disease |  | 3724 (46.3%) | 443 (67.2%) | 3281 (44.5%) |
| **SOCIODEMOGRAPHICS** |  |  |  |  |
| Age | Mean (SD) | 23.4 (17.5) | 29.6 (19.1) | 22.9 (17.3) |
| Sex - Female |  | 4225 (52.6%) | 395 (59.9%) | 3830 (51.9%) |
| Majority tribe |  | 5969 (74.3%) | 508 (77.1%) | 5461 (74.0%) |
| Majority religion |  | 5992 (74.5%) | 488 (74.1%) | 5504 (74.6%) |
| Highest level of education attained | Mean (SD) | 3.44 (2.98) | 3.76 (3.10) | 3.42 (2.97) |
| **Occupation** |  |  |  |  |
| Farmer |  | 1183 (14.7%) | 154 (23.4%) | 1029 (13.9%) |
| Fisherman |  | 610 (7.6%) | 58 (8.8%) | 552 (7.5%) |
| Fishmonger |  | 220 (2.7%) | 31 (4.7%) | 189 (2.6%) |
| None/Other |  | 6025 (75.0%) | 416 (63.1%) | 5609 (76.0%) |
| Home quality score | Mean (SD) | 5.92 (3.49) | 6.00 (3.57) | 5.91 (3.48) |
| HH social status |  | 1079 (13.4%) | 84 (12.7%) | 995 (13.5%) |
| Number of individuals in HH | Mean (SD) | 4.24 (1.80) | 3.84 (1.60) | 4.28 (1.82) |
| Deaths in HH (past 3 yrs) |  | 694 (8.6%) | 74 (11.2%) | 620 (8.4%) |
| Years HH has lived in village | Mean (SD) | 20.5 (15.3) | 20.6 (15.8) | 20.5 (15.3) |
| Home owned |  | 7049 (87.7%) | 566 (85.9%) | 6483 (87.9%) |
| Number of rooms | Mean (SD) | 2.23 (1.19) | 2.23 (1.44) | 2.23 (1.17) |
| **WATER, SANITATION AND HYGIENE (WASH)** |  |  |  |  |
| Improved drinking water source |  | 4464 (55.5%) | 414 (62.8%) | 4050 (54.9%) |
| HH treats drinking water |  | 1791 (22.3%) | 184 (27.9%) | 1607 (21.8%) |
| Improved sanitation facility in home |  | 4830 (60.1%) | 367 (55.7%) | 4463 (60.5%) |
| Basic hygiene facility in home |  | 712 (8.9%) | 102 (15.5%) | 610 (8.3%) |
| **SPATIAL FACTORS** |  |  |  |  |
| Min. dist. (km) to drug shop | Mean (SD) | 1.08 (1.19) | 1.22 (1.24) | 1.07 (1.19) |
| Min. dist. (km) to gov't. health centre | Mean (SD) | 2.65 (1.64) | 2.54 (1.61) | 2.66 (1.64) |
| **District** |  |  |  |  |
| Buliisa |  | 2455 (30.5%) | 151 (22.9%) | 2304 (31.2%) |
| Mayuge |  | 2270 (28.2%) | 192 (29.1%) | 2078 (28.2%) |
| Pakwach |  | 3313 (41.2%) | 316 (48.0%) | 2997 (40.6%) |
| **STUDY DESIGN FACTOR** |  |  |  |  |
| **Year of recruitment** |  |  |  |  |
| 2022 |  | 5298 (65.9%) | 529 (80.3%) | 4769 (64.6%) |
| 2023 |  | 1748 (21.7%) | 104 (15.8%) | 1644 (22.3%) |
| 2024 |  | 992 (12.3%) | 26 (3.9%) | 966 (13.1%) |

There were 8.2% (659/8038) participants who sought any medical care in the month before recruitment. Most participants who sought care visited government health centres (67.5%, 445/659), with 28.4% (187/659) visiting private health clinics, 2.9% (19/659) visiting local medicine shops, 0.9% (6/659) seeking care from VHTs, and only 0.3% (2/659) seeking other types of care including from traditional healers. The percentage of participants who sought care in the past month was 9.5% (316/3313) in Pakwach, 6.2% (151/2455) in Buliisa, and 8.5% (192/2270) in Mayuge (χ^2^ = 21.8, p < 0.001). Among those who sought care, 80.4% (254/316) in Pakwach, 81.5% (123/151) in Buliisa, and 35.4% (68/192) in Mayuge, visited a government health centre excluding approaching VHTs (χ^2^ = 127.4, p < 0.001). Overall, the usual source of care was most frequently a government health centre (84.8%, 1858/2191), followed by private clinics (11.5%, 252/2191), VHTs (2%, 44/2191), and local medicine shops (1.7%, 37/2191). The usual source of care sought by HHs varied significantly by district (χ^2^ = 365.7, p < 0.001). In Buliisa, 92.5% (689/745) HHs reported government health centres as their usual source of care, with a smaller proportion reporting private clinics (6.6%, 49/745), and very few reporting local medicine shops (0.8%, 6/745) or VHTs (0.1%, 1/745). In Pakwach, 89.2% (800/897) relied on government health centres, while local medicine shops (3%, 27/897), private clinics (3.1%, 28/897), and VHTs (4.7%, 42/897) were reported less often. In contrast, in Mayuge only 67.2% (369/549) reported government health centres as their usual source of care, whereas nearly one-third (31.9% 175/549) used private clinics, with very few reporting local medicine shops (0.7%, 4/549) or VHTs (0.2%, 1/549).

Self-reported medical history including history of IDs, history of NCDs, and history of any disease were weakly correlated with seeking care (Spearman’s ρ = 0.122, p < 0.001; ρ = 0.039, p< 0.001; ρ = 0.125, p< 0.001). Among those who sought care, selfreported medical history was not significantly associated with seeking care at a PHC centre. Participant characteristics including biomedical factors, sociodemographics, HH WASH, and spatial factors are summarised in Table 1.

### Distance to PHC centres versus local medicine shops

Across 2191 HHs, the median distance to the nearest local medicine shop was 0.46 km (IQR 0.15-2.07), and for PHC centres the median distance was 2.4 km (IQR 1.47-3.95). Overall, 99.5% (2179/2191) of HHs were located within five km of a medicine shop, and 88.5% (1939/2191) were within five km of a PHC centre (Supplementary Figure S1). The difference in HH distances to PHC centres versus local medicine shops was significant overall and within each district (Wilcoxon rank-sum test p < 0.001; Supplementary Table S4). Among those who sought care at a government health centre in the past month, 20.4% (91/445) reported a travel time of within 10 minutes, 26.3% (117/445) within 30 minutes, 18.2% (81/445) reported a travel time of within 60 minutes, 15.7% (70/445) 1-2 hours, 17.5% (78/445) more than two hours, and 1.8% (8/445) did not know their travel times. However, those who reported a travel time of more than one hour had a significantly higher median distance to a government health facility than those who reported a travel time of more than two hours (Table 2). Among the 10 government health centres recorded in our study, there were six of level Health Centre II (HCII) and four of level Health Centre III (HCIII), with one HCII and two HCIIIs in Pakwach, two HCIIs and one HCIII in Buliisa, and two HCIIs and two HCIIIs in Mayuge.

**Table 2.** Distances (km) to government health centres by travel time. Obs. 445. Travel time was collected on an individual level, while minimum distances were calculated by HH. As a result, HHs may have appeared more than once in the calculation of these medians and IQRs.

| Travel time | Median | IQR |
| --- | --- | --- |
| Within 10 minutes | 0.87 | 0.52-2.78 |
| Within 30 minutes | 1.96 | 0.96-2.91 |
| Within 60 minutes | 2.01 | 1.61-3.73 |
| 1-2 hours | 3.89 | 2.06-4.04 |
| More than 2 hours | 2.10 | 1.8-3.83 |
| Do not know | 1.86 | 1.41-1.9 |

### Final diagnoses and costs

There was little variation in the diagnoses assigned to participants at government health centres, while costs of care varied. Among participants who sought care from a government health centre and had a recorded final diagnosis (76.63%, 341/445), the most common diagnosis was malaria (57.18%, 195/341) (Supplementary Table S5). Among participants who visited a government health centre (excluding those seeking care from VHTs), 36.9% (164/445) had an in-patient stay. Only 34.9% (230/659) reported to receive free care (or to incur no costs excluding transport costs) from government health centres or VHTs. For the paid care received by 65.9% (429/659) of participants, the median amount paid was 10000 UGX or approximately 2.5 USD (IQR 4000-30000). Among participants who had malaria in their final diagnoses, only 45.1% (88/195) reported no cost of care, and others reported a median cost of 10000 Ugandan Shillings (IQR 5000-20000). Among participants who reported having stayed at a government health centre for in-patient care, 47.6% (78/164) reported no cost of care, and others reported a median cost of 24500 Ugandan Shillings (IQR 8000-53750).

### Biomedical, socioeconomic, and spatial disparities

Figure 1 shows the multilevel model for any care sought for all participants with random effects at the village and HH levels. Among all 8038 participants, a history of any disease and the presence of symptoms in the month preceding the study were associated with higher odds of seeking care. Among the study participants, those reporting any symptoms (OR = 2.85, 95% CI: 1.87–4.34), having a prior diagnosis of an infectious or non-communicable disease (OR = 2.06, 95% CI: 1.64–2.59), older age per five-year increase (OR = 1.1, 95% CI: 1.08–1.13), and being female (OR = 1.39, 95% CI: 1.15–1.68) had higher odds of seeking care. Care-seeking was lower among those recruited in 2024 compared to the first study participants recruited in 2022, with differences between participants in 2022 and 2023 being borderline significant based on the FAR estimates (95% CI: 0.37–0.96). The empty multilevel model had an intraclass correlation coefficient (ICC) of 0.261 at the HH level and 0.138 at the village level. After adjustment, the ICCs were 0.277 at the HH level and 0.100 at the village level. In adults aged 20 and above, associations were similar to the overall model with the exception of age, female gender, number of individuals in HH and improved drinking water source not being selected into the model (Supplementary Figure S2).

**Fig. 1.**
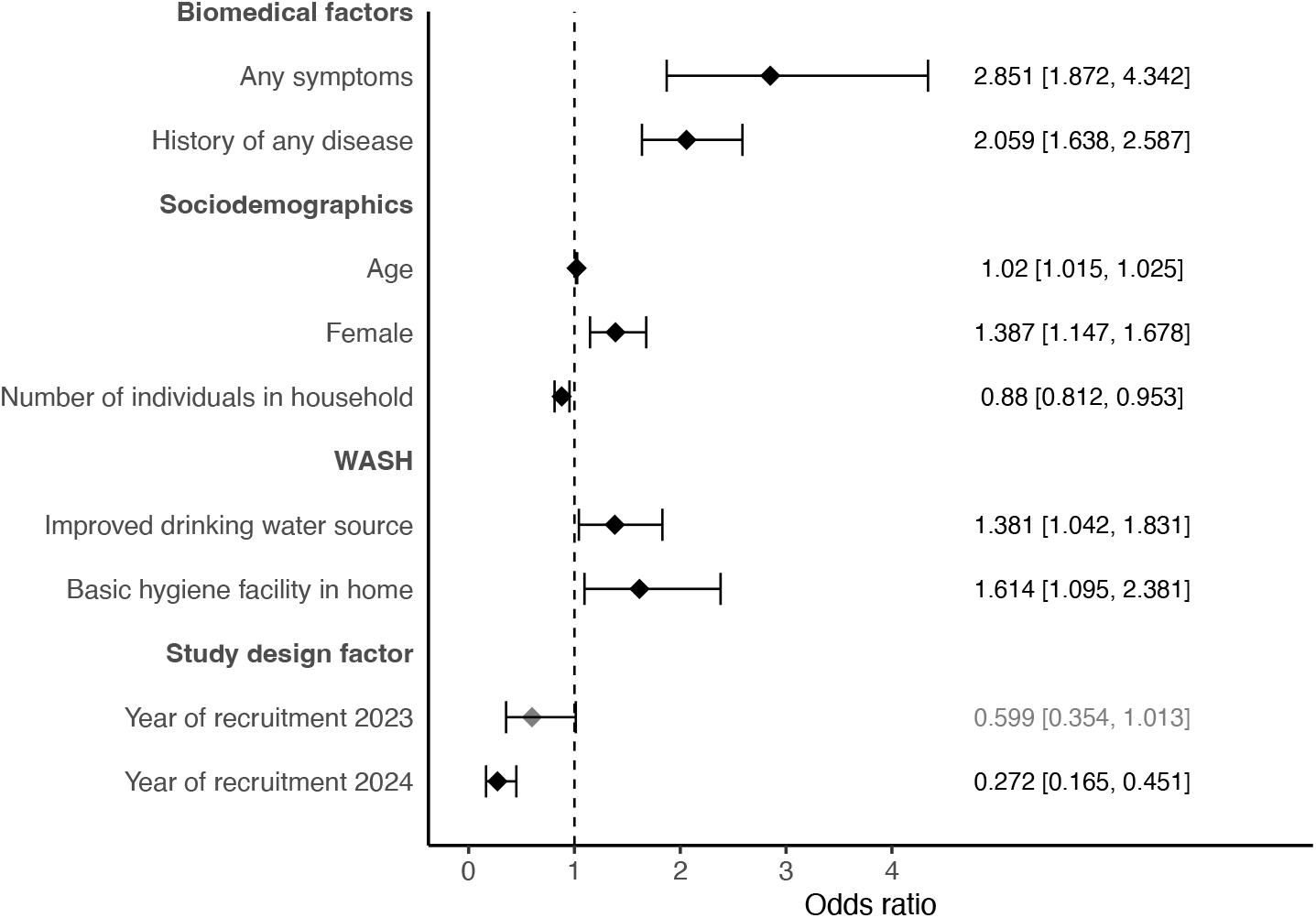
Model for any care sought. Multilevel model for any care sought for all participants (*N* = 8038) with random effects at the village and HH levels. VIFs*<*10 for all covariates, and AUC for 10-fold cross-validation was 0.714. Participants recruited in the year 2024 had significantly lower odds of seeking care, with a FAR of 0.27 (CI 0.17-0.43) compared to the reference year. Similarly, participants recruited in 2023 also exhibited significantly lower care-seeking odds, with a FAR of 0.6 (CI 0.37-0.96).

Among those who sought care, region/district was a key determinant of whether care was obtained from government health centres and VHTs (government care) versus privately (Figure 2). Participants from the Western districts of Buliisa and Pakwach had similarly higher odds of seeking government care compared to those from Mayuge, at 9.40 and 10.77 times higher, respectively. A higher home quality score was only marginally positively associated with higher odds of seeking government care. Having an improved drinking water source in the home was consistently positively associated with seeking government care (as well as any care) while having improved sanitation within the home was negatively associated with seeking government care. When reanalysed for adults only (Supplementary Figure S3), home quality score was no longer relevant and current alcohol consumption was significant while all other variables remained unchanged. Current alcohol consumption was associated with lower odds of seeking government care (OR = 0.43, 95% CI 0.24–0.79) compared with adults who did not drink alcohol.

**Fig. 2.**
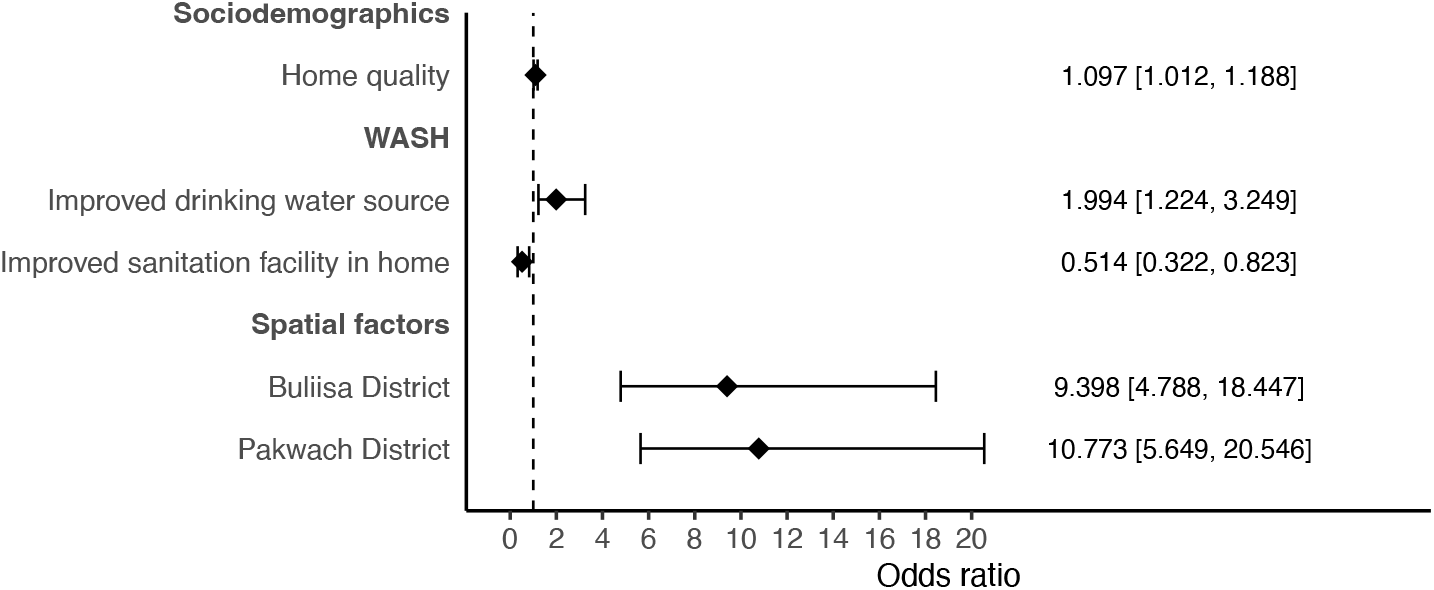
Model for type of care sought. Logistic regression model for the type of care sought for participants who sought care (*n* = 659) with 95% confidence intervals calculated using clustered standard errors at the HH level. VIFs*<*10 for all covariates, and AUC for 10-fold cross-validation was 0.765.

Figure 3 shows the model predicting the usual source of care (government health centre and VHTs versus private health care). Participants with an improved drinking water source in the home had higher odds of using government care as their usual source of care (OR = 1.76, 95% CI: 1.33–2.35) than participants without access to an improved drinking water source. Belonging to a home that was further from a government health centre was negatively associated with having government care as the usual source of care. For each one km further a HH was from a government health centre, the HH had lower odds (OR = 0.72, 95% CI: 0.66–0.79) of having government care as their usual source of care. As with the models for any care sought and the type of care sought, Western Districts of Pakwach and Buliisa were more likely to have the usual source of care be government care when compared to Mayuge (Odds ratios 4.70-5.64). Later years of study recruitment (2023 and 2024) compared to 2022 were positively associated with reporting government care as the usual source of care (FAR = 5.55, 95% CI: 2.96–10.42 for 2023; FAR = 4.35, 95% CI: 0.37–0.96 for 2024).

**Fig. 3.**
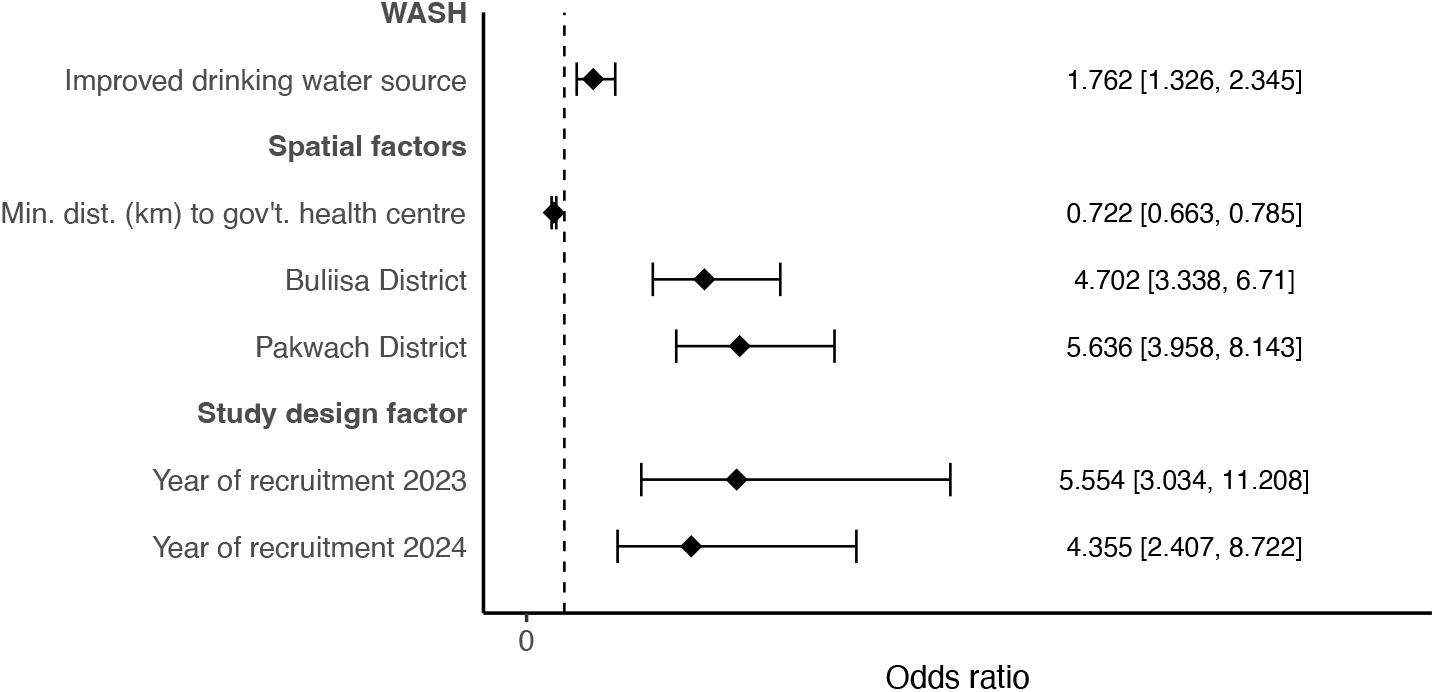
Model for usual source of care sought by HH. Logistic regression model for the type of care sought for participants who sought care (*N* = 2191). VIFs*<*10 for all covariates, and AUC for 10-fold cross-validation was 0.802. Participants recruited in the years 2023 and 2024 had significantly lower odds of seeking care, with FARs of 5.55 (CI 2.96-10.42) and 4.35 (CI 2.34-8.09), compared to the reference year.

## Discussion

Access and utilisation of PHC services remain a challenge in rural areas of sub-Saharan Africa. We conducted a large-scale study within the SchistoTrack cohort on the access and utilisation of PHC services for 8038 participants aged one year and older in three rural districts of Uganda. A comprehensive set of variables was examined that related to medical, socioeconomic, and geographic influences on the decisions to access any care, the type of care chosen, and the usual source of care. The usage of PHC services was low at 67.5% of participants who sought care within the month preceding the study, and was influenced by several complex medical and social factors beyond conventionally studied barriers of the distance of a HH to a PHC centre.

Being currently ill (with symptoms in the past one month) or having a past diagnosis by a health worker of any illness positively influenced the odds of seeking any care. Individuals must recognise or prioritise a problem before seeking care. Past research has suggested that more complex cases of perceived illness, focused on symptom reporting in rural Uganda, are likely to be handled in private facilities or with traditional healers [24]. As this study only separated IDs from NCDs, it was not possible to understand how disease complexity influenced the choice of seeking any care or the type of care. Future work is needed to map the medical history of participants observed in this study to international disease ontologies and assign a disease severity, treatment complexity, or prognosis rating. Recent qualitative evidence from PHC facility use for NCDs in low- and middle-income countries suggests that barriers such as low awareness, limited availability of medicines, and weak continuity of care may further affect care-seeking decisions [33]. Despite a growing burden of IDs and NCDs in rural sub-Saharan Africa and the use of NCDs to signify illnesses requiring routine care as litmus tests for functioning PHC systems, we found no difference in the odds of seeking PHC based on the history of IDs or NCDs [3]. These results suggest that individuals are still likely to go to a health facility for ID care, despite widespread community-based treatment programmes.

The most common diagnosis received from a PHC centre for a participant seeking care was malaria. Given we focused on care seeking within the one month preceding the study, it is possible that the diagnoses reflect the true burden of disease in our study areas. Participants were recruited at the beginning of the dry season when malaria prevalence is expected to peak following a long period of exposure to infected mosquitoes when mosquito abundance is highest in the preceding rainy season [34]. Alternatively, our areas have been shown to be hyperendemic with malaria, with over 41% of participants aged one year and older estimated to be infected, predominantly with *Plasmodium falciparum* [35]. This endemicity may result in most participants having some baseline level of parasitemia that would always show positive on a malaria rapid diagnostic test even without symptoms or obvious clinical implications. In this case, malaria may be over diagnosed when the underlying illness or cause that is resulting in an individual seeking care remains unaddressed. To understand whether malaria is overburdening PHC centres, there is a need for future work to assess how the medical reason for seeking care matches the final diagnosis given (focusing on patient priorities) and to account for multimorbidity where diverse causes beyond malaria may give rise to similar or interacting clinical presentations.

Current alcohol consumption among adults aged 20 years and older was associated with lower odds of seeking any care and, among those who sought care, seeking PHC. Reasons for lower care-seeking behaviour are likely complex. Current drinkers have been shown to have accompanying mental health illnesses [36]. There may also be stigma associated with medical problems due to alcohol use that may prevent someone from seeking care or trusting PHC [37]. Many heavy alcohol users perceive health services as unhelpful for their condition, and those who have sought care often feel their alcohol use was overlooked, discouraging further engagement [38]. Alternatively, alcohol use might generally affect individuals’ inclination towards preventative health measures. Other variables of preventative health behaviour, such as having access to improved drinking water sources were positively associated with seeking any care, seeking PHC among those who sought care, and having PHC as the usual source of care. However, the effect of alcohol use is likely also gender-dependent, as most current drinkers in SchistoTrack are male [39]. Regardless, overall, there is a need for future studies to explore the role of health outreach campaigns for current drinkers by focusing on increased sensitisation and access to care, improved awareness among health care workers, and potentially educational campaigns on preventative health.

Simple demographics were able to profile participants who were less likely to seek any care, but did not influence the choice of the type of care sought. Females, older participants, and participants belonging to smaller HHs (with fewer members) had higher odds of seeking any care within the month preceding this study. It is unknown whether being female and older is an indication of being more likely to need care or whether females are responsible for the medical decisions of the HH and may be a source of information for other HH members on care-seeking decisions. It could be possible that this role might result in increased awareness or preventative health inclinations among this group. Females who are given increased autonomy in intra-HH decision-making have been shown to have increased engagement with PHC services for antenatal care in Uganda [37]. Previous research has also shown that females are more likely to seek care for malaria in Uganda, and malaria was the most common diagnosis given at PHC centres, and also suggesting that females have a higher tendency to seek care than males [40]. Age was only relevant in models that included children, hence indicating that it was not the elderly among the adults who had higher odds of seeking care, e.g., due to frailty. Instead, there was a discrepancy in seeking care between children and adults. Our study only considered older children from 5-17 years of age, hence not capturing potentially high rates of illness due to diarrhoea or other common causes in children under five years. Older children may not be brought to care either because they are less likely to fall ill compared to adults or potentially due to having access to health campaigns within the communities. For HH size, larger HHs may have fewer resources (if the HH is large due to having many unemployed or in-school children) and potentially less available funds to support transport costs, given all care seeking in our areas did not involve traditional healers within the village. Overall, our study suggests that demographic factors highlight inequalities in access to any type of care for a participant, and future research is needed to determine whether this is an active choice of the participant/HH, due to social or infrastructural barriers, or influenced by social norms/expectations.

There were higher odds of a HH having PHC as their usual source of care the closer they lived to a PHC centre. Distance to health care faciltiies is a well-established influence of health care access. The Uganda Ministry of Health strives to have 100% of HHs within five km walking distance of a Health Centre III, which is a PHC centre that at minimum should have an outpatient department and maternity ward [41]. However, in our study only 88.5% of HHs were within a five km catchment of any PHC centre, including even Health Centre II facilities that only provide basic outpatient care such as the distribution of antimalarials or antenatal checks. In our study, the participants furthest from a facility did not have the greatest travel times to the facility, suggesting there are thresholds at which participants need to pay for transport instead of walking. Uganda is currently building 398 more Health Centre IIIs across the country [42]. However, reducing the distance to HHs is likely not the only factor that will affect care access. We observed stark regional disparities where our Western study districts of Pakwach and Buliisa had higher odds of using PHC services either by accessing PHC within the month preceding the study or having PHC as their usual source of care. Infrastructure in our Eastern district of Uganda is extensively developed with many private hospitals, local medicine shops, and also more government health centres at all levels from HCII to HCIV than our Western districts [27]. This extensive and high-quality infrastructure provides more choice to the residents. And, it may be that there is a preference (and the financial resources from fishing, which may be more commercialised in the Eastern district) to enable participants to choose to use private sources. However, there was less reporting of private medicine shop use in Mayuge, which might be attributed to stronger regulatory enforcement by the National Drug Authority, given its proximity to the capital city Kampala. Alternatively, future work would be needed to assess whether participants in the Eastern district of Uganda have less trust of government services. Regardless, simply building more government health centres would not necessarily change individual choices of where to receive care and there is a need to understand community needs, preferences, and social beliefs as well as other diverse operational levers of PHC strengthening [23].

The health system in Uganda, beyond transportation and other indirectly incurred costs outside of the facilities, by policy is meant to be free-of-cost to patients visiting government-funded facilities. Yet, in our study, most participants who sought care from government facilities, including even routine care for malaria, incurred out-of-pocket expenses as has been previously documented elsewhere [43]. These expenses may be due to government facilities being out-of-stock of prescribed medicines or tests being unavailable within the facility; both potentially resulting in patients being sent to private facilities. Considering how to improve supply chains and cost-share with patients is important for ensuring affordability of PHC to progress towards UHC.

The strengths in our study lie in the comprehensive of covariates studied, diverse health access outcomes, large-scale rural population studied, and models of good to high predictive accuracy. Some limitations include that we focus on the main source of care either within the past month or usual source used by the HH. However, care continuity and care seeking is complex, often with individuals seeking care from many sources and we have not captured here the sequence at which individuals sought care (e.g. first going to a government facility then a private centre). More information also is needed on the access to informal care in the event it was underreported in our study, again possibly due to not capturing the sequence of care. The year of recruitment was an important influence on the use of PHC as the usual source of care. The study may have positively influenced participants who were recruited in later years given that SchistoTrack, in collaboration with the Uganda Minsitry of Health, engages local government health workers to join the central Kampala teams to clinically assess participants. It may be that participants were able to build more trust in the local health system as they observed the health workers from the government centres working within their villages. Regardless, this influence opens avenues for future research studies to strengthen participants’ connections to government health systems through capacity building and local employment of teams.

In conclusion, we have shown that the access to health care and usage of PHC in rural sub-African settings is determined by a complex set of factors. Medical, socioeconomic, and geographic disparities remain and influence the usage of PHC, potentially undermining goals of UHC. There are key differences in the access to any care and the choice of using PHC where demographics should be targeted for the former issue and medical and geographic disparities should be the focus of the latter issue. If replicated in other countries, we anticipate our findings can guide strategies for improving the access to and utilisation of PHC services in sub-Saharan Africa.

## Declarations

## Acknowledgments

We are thankful to the SchistoTrack Group for extensive feedback, especially Fabian Reitzug and Max Lang for early discussions. We also thank Seun Anjorin for input on the initial construction of the medical history variables. Many thanks especially go to the SchistoTrack teams in Uganda, in particular the HH surveyors, district focal people, VHTs, and the study participants.

## Data availability

The datasets generated and/or analysed for the current study are not available due to the ongoing nature of the cohort and the privacy agreements and ethics procedures in place with participants.

## Code availability

No new model code was developed.

## Materials availability

Not applicable.

## Ethics approvals

Written informed consent was provided by participants (or their guardians for children). The study was approved by Oxford Tropical Research Ethics Committee OxTREC 509-21; Vector Control Division Uganda Ministry of Health VCDREC146; and Uganda National Council for Science and Technology UNCST HS 1664ES.

## Consent for publication

Not applicable.

## Author contributions

GFC accepts full responsibility for the finished work and/or the conduct of the study, had access to the data, and controlled the decision to publish. Conceptualisation: GFC. Data curation: CP, YCZ, BT, BN, NBK, GFC. Formal analysis: SM, CP. Funding acquisition: GFC. Investigation: SM, CP, GFC. Methodology: SM, CP, GFC. Project administration: NBK, GFC. Resources: NBK, GFC. Software: GFC. Supervision: GFC. Validation: CP. Visualization: CP. Writing – original draft: SM, CP, GFC. Writing – review & editing: SM, CP, BN, BT, YCZ, CKO, NBK, GFC.

