## Supplementary material for "Medical, socioeconomic, and geographic disparities in primary health care access and utilization: A population-based study of 8038 individuals aged one year and older in rural Uganda"

### Supplementary information

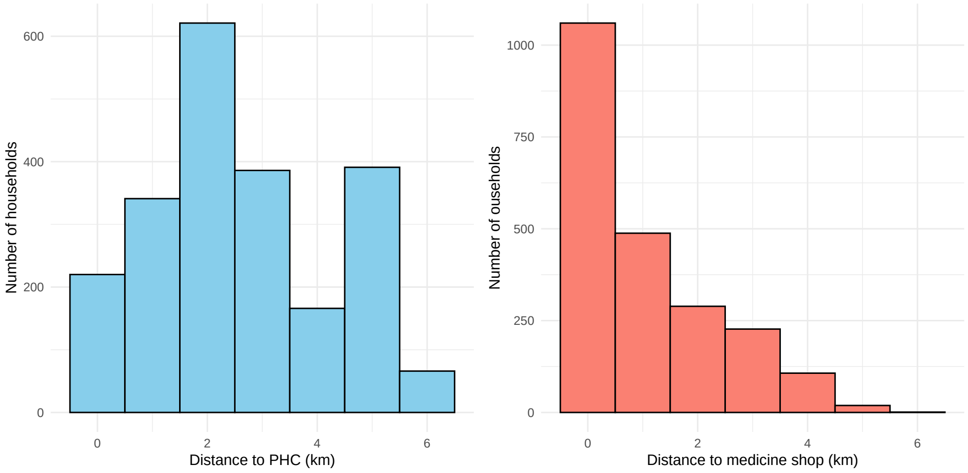


**Fig. S1** Histogram of HH distances to PHCs and medicine shops.

**Table S1** Self-reported medical history.

| | Total | Care seeking | Not care seeking |
|---|---|---|---|
| | (N=8038) | (N=659) | (N=7379) |
| **HISTORY OF INFECTIOUS DISEASES** | | | |
| **Uncomplicated malaria** | 1642 (20.4%) | 216 (32.8%) | 1426 (19.3%) |
| **Complicated malaria** | 507 (6.3%) | 84 (12.7%) | 423 (5.7%) |
| **Encephalitis** | 108 (1.3%) | 15 (2.3%) | 93 (1.3%) |
| **Pneumonia** | 207 (2.6%) | 36 (5.5%) | 171 (2.3%) |
| **Pertussis** | 679 (8.4%) | 73 (11.1%) | 606 (8.2%) |
| **Scabies** | 135 (1.7%) | 19 (2.9%) | 116 (1.6%) |
| **Tetanus** | 33 (0.4%) | 2 (0.3%) | 31 (0.4%) |
| **Typhoid** | 1306 (16.2%) | 165 (25.0%) | 1141 (15.5%) |
| **Cholera** | 625 (7.8%) | 63 (9.6%) | 562 (7.6%) |
| **Dysentery** | 339 (4.2%) | 45 (6.8%) | 294 (4.0%) |
| **Diarrhoea** | 608 (7.6%) | 65 (9.9%) | 543 (7.4%) |
| **Tuberculosis** | 65 (0.8%) | 17 (2.6%) | 48 (0.7%) |
| **Meningitis** | 33 (0.4%) | 7 (1.1%) | 26 (0.4%) |
| **Hepatitis B** | 41 (0.5%) | 5 (0.8%) | 36 (0.5%) |
| **Hepatitis C** | 17 (0.2%) | 2 (0.3%) | 15 (0.2%) |
| **Measles** | 45 (0.6%) | 12 (1.8%) | 33 (0.4%) |
| **HIV** | 127 (1.6%) | 20 (3.0%) | 107 (1.5%) |
| **Sepsis** | 62 (0.8%) | 6 (0.9%) | 56 (0.8%) |
| **Leprosy** | 28 (0.3%) | 2 (0.3%) | 26 (0.4%) |
| **Brucellosis** | 27 (0.3%) | 4 (0.6%) | 23 (0.3%) |
| **Gut worms** | 952 (11.8%) | 129 (19.6%) | 823 (11.2%) |
| **Bilharzia** | 330 (4.1%) | 58 (8.8%) | 272 (3.7%) |
| **Trachoma** | 285 (3.5%) | 47 (7.1%) | 238 (3.2%) |
| **Hemorrhagic fever** | 49 (0.6%) | 6 (0.9%) | 43 (0.6%) |
| **COVID** | 19 (0.2%) | 2 (0.3%) | 17 (0.2%) |
| **HISTORY OF NON-COMMUNICABLE DISEASES** | | | |
| **Epilepsy** | 55 (0.7%) | 8 (1.2%) | 47 (0.6%) |
| **Sickle cell disease** | 49 (0.6%) | 9 (1.4%) | 40 (0.5%) |
| **Anaemia** | 93 (1.2%) | 14 (2.1%) | 79 (1.1%) |
| **Liver disease** | 180 (2.2%) | 25 (3.8%) | 155 (2.1%) |
| **Kidney disease** | 58 (0.7%) | 13 (2.0%) | 45 (0.6%) |
| **Asthma** | 152 (1.9%) | 19 (2.9%) | 133 (1.8%) |
| **Eczema** | 752 (9.4%) | 68 (10.3%) | 684 (9.3%) |
| **Cancer** | 41 (0.5%) | 6 (0.9%) | 35 (0.5%) |
| **Depression** | 102 (1.3%) | 22 (3.3%) | 80 (1.1%) |
| **Anxiety** | 82 (1.0%) | 13 (2.0%) | 69 (0.9%) |
| **Psychosis** | 17 (0.2%) | 3 (0.5%) | 14 (0.2%) |

**Table S2** HH characteristics.

| | Total | HH health care other | HH health care PHC |
|---|---|---|---|
| | (N=2191) | (N=289) | (N=1902) |
| **HH HEALTH** | | | |
| **History of IDs** | 1470 (67.1%) | 194 (67.1%) | 1276 (67.1%) |
| **History of NCDs** | 895 (40.8%) | 103 (35.6%) | 792 (41.6%) |
| **History of any disease** | 1601 (73.1%) | 213 (73.7%) | 1388 (73.0%) |
| **SOCIODEMOGAPHICS** | | | |
| **Highest level of education attained of HH head** | | | |
| Mean (SD) | 4.66 (3.31) | 4.90 (3.20) | 4.62 (3.33) |
| **Occupation of HH head** | | | |
| Farmer | 571 (26.1%) | 85 (29.4%) | 486 (25.6%) |
| Fisherman | 541 (24.7%) | 81 (28.0%) | 460 (24.2%) |
| Fishmonger | 107 (4.9%) | 9 (3.1%) | 98 (5.2%) |
| None/Other | 972 (44.4%) | 114 (39.4%) | 858 (45.1%) |
| **Home quality score** | | | |
| Mean (SD) | 5.76 (3.44) | 7.37 (3.58) | 5.52 (3.35) |
| **HH social status** | 257 (11.7%) | 24 (8.3%) | 233 (12.3%) |
| **Number of individuals in HH** | | | |
| Mean (SD) | 3.66 (1.46) | 3.92 (1.40) | 3.62 (1.46) |
| **Deaths in HH (past 3 yrs)** | 161 (7.3%) | 19 (6.6%) | 142 (7.5%) |
| **Years HH has lived in village** | | | |
| Mean (SD) | 20.0 (15.6) | 20.3 (14.4) | 20.0 (15.8) |
| **Home owned** | 1898 (86.6%) | 252 (87.2%) | 1646 (86.5%) |
| **Number of rooms** | | | |
| Mean (SD) | 2.14 (1.17) | 2.17 (1.06) | 2.14 (1.18) |
| **WATER, SANITATION AND HYGIENE (WASH)** | | | |
| **Improved drinking water source** | 1184 (54.0%) | 141 (48.8%) | 1043 (54.8%) |
| **HH treats drinking water** | 489 (22.3%) | 74 (25.6%) | 415 (21.8%) |
| **Improved sanitation facility in home** | 1253 (57.2%) | 192 (66.4%) | 1061 (55.8%) |
| **Basic hygiene facility in home** | 183 (8.4%) | 32 (11.1%) | 151 (7.9%) |
| **SPATIAL FACTORS** | | | |
| **Min. dist. (km) to drug shop** | | | |
| Mean (SD) | 1.11 (1.22) | 0.618 (0.912) | 1.19 (1.24) |
| **Min. dist. (km) to gov't. health centre** | | | |
| Mean (SD) | 2.62 (1.60) | 3.34 (1.68) | 2.51 (1.56) |
| **District** | | | |
| Buliisa | 745 (34.0%) | 55 (19.0%) | 690 (36.3%) |
| Mayuge | 549 (25.1%) | 179 (61.9%) | 370 (19.5%) |
| Pakwach | 897 (40.9%) | 55 (19.0%) | 842 (44.3%) |
| **STUDY DESIGN FACTOR** | | | |
| **Year of recruitment** | | | |
| 2022 | 1459 (66.6%) | 267 (92.4%) | 1192 (62.7%) |
| 2023 | 493 (22.5%) | 11 (3.8%) | 482 (25.3%) |
| 2024 | 239 (10.9%) | 11 (3.8%) | 228 (12.0%) |

**Table S3** Characteristics of adults.

| | Total | Care seeking | Not care seeking |
|---|---|---|---|
| | (N=3780) | (N=420) | (N=3360) |
| **BIOMEDICAL FACTORS** | | | |
| **Any symptoms** | 232 (6.1%) | 59 (14.0%) | 173 (5.1%) |
| **IDs for other HH members** | 2299 (60.8%) | 295 (70.2%) | 2004 (59.6%) |
| **NCDs for other HH members** | 1210 (32.0%) | 151 (36.0%) | 1059 (31.5%) |
| **History of IDs** | 1880 (49.7%) | 265 (63.1%) | 1615 (48.1%) |
| **History of NCDs** | 944 (25.0%) | 151 (36.0%) | 793 (23.6%) |
| **History of any disease** | 2094 (55.4%) | 305 (72.6%) | 1789 (53.2%) |
| **Current alcohol consumption** | 616 (16.3%) | 76 (18.1%) | 540 (16.1%) |
| **Current smoker** | 445 (11.8%) | 51 (12.1%) | 394 (11.7%) |
| **SOCIODEMOGAPHICS** | | | |
| **Age** | | | |
| Mean (SD) | 38.9 (13.4) | 40.9 (14.5) | 38.7 (13.2) |
| **Sex - Female** | 2131 (56.4%) | 256 (61.0%) | 1875 (55.8%) |
| **Majority tribe** | 2838 (75.1%) | 331 (78.8%) | 2507 (74.6%) |
| **Majority religion** | 2796 (74.0%) | 312 (74.3%) | 2484 (73.9%) |
| **Highest level of education attained** | | | |
| Mean (SD) | 4.57 (3.26) | 4.41 (3.23) | 4.59 (3.26) |
| **Occupation** | | | |
| Farmer | 1108 (29.3%) | 137 (32.6%) | 971 (28.9%) |
| Fisherman | 589 (15.6%) | 58 (13.8%) | 531 (15.8%) |
| Fishmonger | 218 (5.8%) | 30 (7.1%) | 188 (5.6%) |
| None/Other | 1865 (49.3%) | 195 (46.4%) | 1670 (49.7%) |
| **Home quality score** | | | |
| Mean (SD) | 5.88 (3.49) | 5.90 (3.55) | 5.88 (3.48) |
| **HH social status** | 498 (13.2%) | 50 (11.9%) | 448 (13.3%) |
| **Number of individuals in HH** | | | |
| Mean (SD) | 3.94 (1.56) | 3.77 (1.66) | 3.96 (1.54) |
| **Deaths in HH (past 3 yrs)** | 299 (7.9%) | 52 (12.4%) | 247 (7.4%) |
| **Years HH has lived in village** | | | |
| Mean (SD) | 20.2 (15.4) | 21.5 (15.6) | 20.0 (15.4) |
| **Home owned** | 3295 (87.2%) | 363 (86.4%) | 2932 (87.3%) |
| **Number of rooms** | | | |
| Mean (SD) | 2.21 (1.22) | 2.25 (1.44) | 2.21 (1.18) |
| **WATER, SANITATION AND HYGIENE (WASH)** | | | |
| **Improved drinking water source** | 2059 (54.5%) | 253 (60.2%) | 1806 (53.8%) |
| **HH treats drinking water** | 863 (22.8%) | 120 (28.6%) | 743 (22.1%) |
| **Improved sanitation facility in home** | 2227 (58.9%) | 227 (54.0%) | 2000 (59.5%) |
| **Basic hygiene facility in home** | 346 (9.2%) | 61 (14.5%) | 285 (8.5%) |
| **SPATIAL FACTORS** | | | |
| **Min. dist. (km) to drug shop** | | | |
| Mean (SD) | 1.09 (1.20) | 1.26 (1.23) | 1.07 (1.19) |
| **Min. dist. (km) to gov't. health centre** | | | |
| Mean (SD) | 2.65 (1.62) | 2.63 (1.63) | 2.65 (1.61) |
| **District** | | | |
| Buliisa | 1225 (32.4%) | 94 (22.4%) | 1131 (33.7%) |
| Mayuge | 1004 (26.6%) | 116 (27.6%) | 888 (26.4%) |
| Pakwach | 1551 (41.0%) | 210 (50.0%) | 1341 (39.9%) |
| **STUDY DESIGN FACTOR** | | | |
| **Year of recruitment** | | | |
| 2022 | 2538 (67.1%) | 324 (77.1%) | 2214 (65.9%) |
| 2023 | 823 (21.8%) | 78 (18.6%) | 745 (22.2%) |
| 2024 | 419 (11.1%) | 18 (4.3%) | 401 (11.9%) |

**Table S4** HH distances to PHCs and medicine shops by district.

| District | Number of HHs | Median distance to PHC (km) | Percentage of HHs within 5km of a PHC | Median distance to medicine shop (km) | Percentage of HHs within 5km of a medicine shop |
|---|---|---|---|---|---|
| Pakwach | 897 | 2.41 (1.75-4.57) | 87.1 | 0.9 (0.33-2.23) | 100 |
| Buliisa | 745 | 2.5 (0.6-3.02) | 100 | 0.34 (0.13-3.04) | 98.4 |
| Mayuge | 549 | 2.05 (1.26-4.86) | 75.2 | 0.18 (0.08-0.4) | 100 |

**Table S5** Reported final diagnoses from government health centres. The categories are not mutually exclusive as some individuals may have had more than one diagnosis. Percentages were computed against a total of 341 individuals who had non-missing information on final diagnoses from government health centres.

| Symptom | Count | Percentage |
|---|---|---|
| **Malaria** | 195 | 57.18 |
| **Other** | 25 | 7.33 |
| **Pregnancy-related** | 17 | 4.99 |
| **Headache** | 9 | 2.64 |
| **Ulcer** | 8 | 2.35 |
| **Cough** | 7 | 2.05 |
| **HIV** | 7 | 2.05 |
| **Typhoid** | 5 | 1.47 |
| **Abdominal pain** | 5 | 1.47 |
| **Flu** | 4 | 1.17 |
| **Epilepsy** | 3 | 0.88 |
| **AIDS** | 3 | 0.88 |
| **Asthma** | 3 | 0.88 |
| **Tuberculosis** | 3 | 0.88 |
| **Worms** | 3 | 0.88 |
| **Dental problem** | 2 | 0.59 |
| **Kidney failure** | 2 | 0.59 |
| **Diabetes** | 2 | 0.59 |
| **Fever** | 2 | 0.59 |
| **Stomach pain** | 2 | 0.59 |
| **Allergy** | 2 | 0.59 |
| **Sickle cell** | 2 | 0.59 |
| **Syphilis** | 2 | 0.59 |
| **Traumatic brain injury** | 1 | 0.29 |
| **Yellow fever** | 1 | 0.29 |
| **Appendicitis** | 1 | 0.29 |
| **Splenomegaly** | 1 | 0.29 |
| **Mental disorder** | 1 | 0.29 |
| **Vomiting blood** | 1 | 0.29 |
| **Anaemia** | 1 | 0.29 |
| **Hepatitis B** | 1 | 0.29 |
| **Thoracic disease** | 1 | 0.29 |
| **Ophthalmic disease** | 1 | 0.29 |
| **Anemia** | 1 | 0.29 |
| **Dysentry** | 1 | 0.29 |
| **Respiratory tract infections** | 1 | 0.29 |
| **Wounds** | 1 | 0.29 |
| **Skin rashes** | 1 | 0.29 |
| **Back pain** | 1 | 0.29 |
| **Hypertension** | 1 | 0.29 |
| **Heart attack** | 1 | 0.29 |
| **Bilharzia** | 1 | 0.29 |
| **Haematuria** | 1 | 0.29 |
| **Intestinal operation** | 1 | 0.29 |
| **Measles** | 1 | 0.29 |
| **Hernia** | 1 | 0.29 |
| **Liver enlargement** | 1 | 0.29 |
| **Spleen enlargement** | 1 | 0.29 |
| **Uterus problem** | 1 | 0.29 |
| **General health check** | 1 | 0.29 |
| **Chest pain** | 1 | 0.29 |

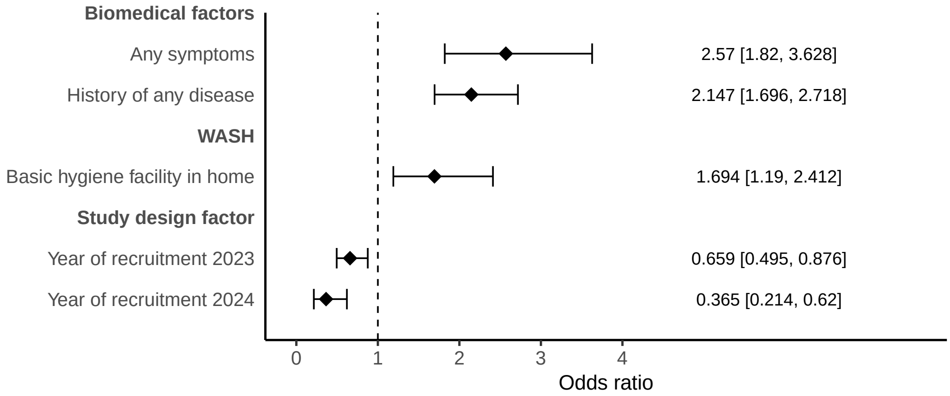


**Fig. S2 Model for any care sought for adults.** Multilevel model for any care sought for individuals aged 20 and above ($n = 3780$) with random effects at the village and HH levels. VIFs$<$10 for all covariates, and AUC for 10-fold cross-validation was 0.656.

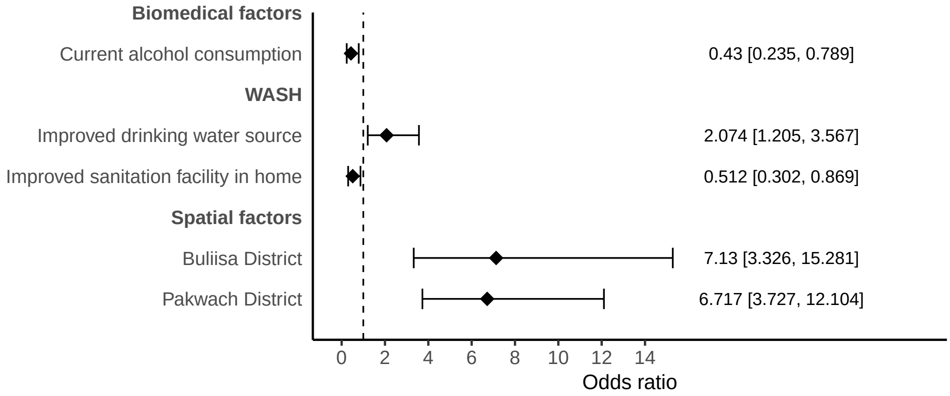


**Fig. S3 Model for type of care sought for adults.** Logistic regression model for the type of care sought for individuals aged 20 and above who sought care ($n = 420$) with 95% confidence intervals calculated using clustered standard errors at the HH level. VIFs$<$10 for all covariates, and AUC for 10-fold cross-validation was 0.745.
